# Longitudinal Adaptive Behavioral Outcomes in Ogden Syndrome by Seizure Status and Therapeutic Intervention

**DOI:** 10.1101/2024.02.23.24303144

**Authors:** Rikhil Makwana, Carolina Christ, Elaine Marchi, Randie Harpell, Gholson J. Lyon

## Abstract

Ogden syndrome, also known as NAA10-related neurodevelopmental syndrome, is a rare genetic condition associated with pathogenic variants in the NAA10 N-terminal acetylation family of proteins. The condition was initially described in 2011, and is characterized by a range of neurologic symptoms, including intellectual disability and seizures, as well as developmental delays, psychiatric symptoms, congenital heart abnormalities, hypotonia and others. Previously published articles have described the etiology and phenotype of Ogden syndrome, mostly with retrospective analyses; herein, we report prospective data concerning its progress over time. Additionally, we describe the nature of seizures in this condition in greater detail, as well as investigate how already-available non-pharmaceutical therapies impact individuals with NAA10-related neurodevelopmental syndrome. Using Vineland-3 scores, we show decline in cognitive function over time in individuals with Ogden syndrome. Sub-domain analysis found the decline to be present across all modalities. Additional investigation between seizure and non-seizure groups showed no significant difference in adaptive behavior outcomes. Therapy investigation showed speech therapy to be the most commonly used therapy by individuals with NAA10-related neurodevelopmental syndrome, followed by occupational and physical therapy. with more severely affected individuals receiving more types of therapy than their less-severe counterparts. Early intervention analysis was only significantly effective for speech therapy, with analyses of all other therapies being non-significant. Our study portrays the decline in cognitive function over time of individuals within our cohort, independent of seizure status and therapies being received, and highlights the urgent need for the development of effective treatments for Ogden syndrome.

## Introduction

N-α-acetyltransferase 10 (NAA10) is the catalytic subunit that, in conjunction with auxiliary subunit NAA15, composes the of N-acetyltransferase A (NatA) complex which functions to acetylate various Ser, Ala, Thr, Gly, Val and Cys residues at the N-terminus of proteins^1^. N-terminal acetylation is a common modification to proteins that has been conserved across species^2–6^ and functions to alter half-life, folding, localization, and expression of various proteins^7–11^. Variations in NAA10 were originally linked with cancer^12–14^, but there has been an increasing amount of evidence suggesting its dysfunction can lead to widespread developmental delays^15–27^. In particular, Ogden Syndrome, also known as NAA10-related neurodevelopmental syndrome, is a primarily X-linked condition associated with pathogenic variants in the NAA10 N-terminal acetylation family of proteins^15^. The condition was first described in 2011 in a family that resided in Ogden, Utah, USA^28,29^. Five males had died in early infancy from a range of cardiac and other defects, all containing a missense change coding for Ser37Pro in the gene NAA10^30^. Since then, other papers have reported additional pathogenic variants in NAA10^15–27^. NAA15, the auxiliary sub-unit in the NatA complex, functions by tethering NAA10 to ribosomes to allow for its co-translational modification function^7,31^. NAA15 variants have also been implicated in developmental diseases including intellectual disability, autism, dystonia, and congenital heart disease^15,32–40^.

The phenotypes for NAA10-related neurodevelopmental syndrome are more variable than those stemming from NAA15 variants possibly due to it having additional function outside of the NatA complex^1,7^. Additionally, the severity depends on the specific pathogenic variant leading to the disease and individual sex as males tend to show more severe symptoms than their female counterparts^15^. Along the spectrum of associated manifestations, there are several cardiac, central nervous system, anatomical, and developmental abnormalities which depend upon the sex and pathogenic variant type for their specific presentation^1,14–20,20–26,28,37,41^. Specific brain abnormalities that have been associated with Ogden syndrome include enlarged ventricles, cerebral dysgenesis, and seizures^15^. Seizures are either focal or generalized areas of excessive and disordered neuronal activity in the brain that lead to symptoms such as myoclonic jerks, hypotonia, hallucinations, abnormal motor posturing and more^42^. Seizures have a prevalence of about .5-1% in the general population with around 150,000 adolescents a year having an unprovoked seizure^43^. In addition to the general distress that unprovoked seizures cause a patient and their caregiver, seizures are associated with developmental delay^44–48^. Seizures have been reported in various cases of NAA10-related neurodevelopmental syndrome^16,19^, however there has not been any definitive links made between the overall incidence of seizure or the incidence of seizure based off a patient’s specific gene pathogenic variant. In writing this paper, we are hoping to uncover any associations between the seizure phenotype and Ogden syndrome, alongside adaptive behavior and the achievement of developmental milestones. In doing so, we hope to identify possible new avenues for treatment, by focusing on identifying and mitigating seizures, to help improve development.

In addition to seizures, individuals with NAA10-related developmental disease often display neurodevelopmental symptoms such as motor and speech delays or even mutism. Intellectual disability is the most prevalent neurologic symptom, affecting 96.8% of the studied cohort^15^. Psychiatric symptoms overlap with autism-like behaviors, where behaviors such as poor eye contact and socialization skills, attention deficits and repetitive behaviors (hand-flapping, tics, echolalia, etc.) are common. Other psychiatric symptoms include harmful and impulsive/compulsive behaviors^15^. There is currently no standard of care that specifically targets these symptoms in individuals with Ogden syndrome. Given the overlap in symptoms between NAA10-related neurodevelopmental syndrome and other neurodevelopmental disorders, we sought to understand how already-available interventions impacted these individuals. The treatments explored in this study include widely used therapies such as speech therapy (also commonly referred to as speech-language therapy), occupational therapy and physical therapy, as well as less traditional therapies including applied-behavior analysis (ABA), equine, water and group therapy.

## Methods

### Participants

The participant population included a group of caregivers and their children that the investigator (G.J.L.) has previously worked with for other research projects. Additional participants were recruited from an online forum (Facebook) where caregivers of children or adults with Ogden syndrome share advice with one another or seek support. Participants were not financially compensated for their time, and they were informed that their participation was completely voluntary and anonymous. Participants were also given the opportunity to receive the results of our analysis as thanks for their participation. Overall, there were a total of 58 participants who completed the survey. Unbeknownst to the authors, one of the study participants shared the survey to the Facebook group composed of other caregivers, some of whom had not previously participated in any of the prior research. This led to 9 individuals who responded to the survey without completing the consent form, sharing their genetic testing results, or having Vineland assessments performed. These data were neither analyzed nor used, and the authors are working to obtain consent from these families in the future. The OS identification numbers, with the key to identify particular research participants, are only known to the study investigators.

### Cognitive Assessment

The individuals diagnosed with Ogden syndrome were given the Vineland Adaptive Behavioral Scales assessment, third edition (Vineland). The Vineland is composed of three core domains, Communication, Daily Living Skills, and Socialization, which are split into three sub-domains and used to assess an individual’s adaptive behavior to determine their levels of personal and social sufficiency^49^. These scores are norm-referenced and the sum of the 3 sub-scores generate an Adaptive behavior composite (ABC) score, providing a more general picture across all domains. In contrast, growth scale values (GSV) are non norm-referenced scores that track performance across test administrations, showcasing progress on an individual level. The Vinelands were administered by two trained assessors at various timepoints in the individuals’ lives with participants ranging in age from less than a year to 40 years of age at the time of the assessment. The caregivers of the participants were the ones who were interviewed and given the Vineland assessment as they knew the participants best and were able to provide the most comprehensive answers.

### Survey

To investigate if there is an association between seizure status and pathogenic variant type, as well as therapy interventions and Vineland-3 score, we surveyed a group of caregivers whose children were diagnosed with Ogden syndrome using a survey administered electronically via Google Forms. The survey was composed of questions asking about the specific types of therapy that the children received, additional information regarding date of start and frequency, as well as the caregivers’ opinions on the therapy. The survey also included questions about if the child experienced seizures, seizure status and type, the pharmaceutical and non-pharmaceutical interventions used for the treatment of symptoms, and general information about the child. Questions were reviewed by the principal investigator and research assistants before being sent out.

### Analysis

#### I. Natural history analysis

Vineland-3 scores were obtained by the PI and associated staff over the course of multiple years, acquiring at least one test per participant. Many participants were able to take it more than once, allowing us to track their performance over time. We conducted a natural history analysis by plotting participants’ ABC scores against their age at the time of the Vineland-3 assessment. This process was repeated for each administration of the test, showcasing their progress over the years. The analysis was filtered further for better visualization of the data, separating participants by gender and pathogenic variant. After plotting each ABC score, participant standard scores and growth scale values in each of the primary domains and sub-domains were also analyzed and then filtered by gender and pathogenic variant to facilitate visualization. The domains of interest were Communication (Com), Daily Living Skills (dls), Socialization (soc), and Motor Skills (mot). The communications subdomains of interest were Receptive (rec), Expressive (exp), and Written (wrn). The daily living skills subdomains of interest were Personal (per), Domestic (dom), and Community (cmm). The socialization subdomains of interest were Play and Leisure (pla), Interpersonal Relationships (ipr), and Coping Skills (cop). The motor subdomains of interest were Gross Motor (gmo) and Fine Motor (fmo). Participant’s sub-scores were tracked over time and graphed, allowing for a more comprehensive view of our cohort’s skills. Motor scores were unable to be collected for individuals with the p.Ala104Asp, p.His16Pro, p.His120Pro, and p.Tyr043Ser pathogenic variants due to differences in the protocol at the time of collecting data. The graphs were further divided by gender and pathogenic variant. When calculating the current snapshot of Vineland scores, all available data points were included, e.g. if a child was administered the Vineland three times, each of the three scores they achieved was included in the average calculations. Ages used were ages at the time of assessment.

#### II. Therapy analysis

The survey requested specific information regarding 8 different types of therapy: Speech, Physical, Occupational, Equine, Water, ABA and Group Therapy, with ‘Other’ being an open-ended option where caregivers could include other non-pharmaceutical forms of intervention. Survey questions included information regarding the date of the participant’s first session, last session if applicable, frequency such as days per week and length per session, whether the caregiver believed the intervention was helpful and why. Information from the survey was summarized to acquire a general picture of the current types of therapies being used by individuals with NAA10-related neurodevelopmental syndrome and the reported parental satisfaction of each. The total number of therapy interventions per individual was compared to their respective ABC score to evaluate whether the number of therapies being received impacted participant’s adaptive behavior. Participants’ ABC scores were further analyzed with the age (in months) they began receiving therapies they reported participating in. This analysis aimed to investigate whether earlier intervention had an impact on participants’ scores. Correlation and linear regression analysis were performed using GraphPad prism with a presumed significance of p<.05 to investigate whether there was a relationship between the variables chosen.

#### III. Seizure analysis

After closing the survey, data was collected and organized in Microsoft Excel. A two-tailed unequal variance t-tests were performed with a presumed p-value of <.05 to determine if there was a difference in ABC standard score or a difference between when developmental milestones were achieved between those with and without seizures. After initial comparisons between the Vineland ABC standard scores were completed, additional analysis was performed on each of the Vineland domains and sub-domains. The data was then input into Prism GraphPad for visualization. Ages used in the analysis were the ages at the time of which the participants caregiver took the Vineland assessment. The Vineland scores used were from the most recent administration. Only participants that had both Vineland data and filled out the survey were used for the analysis; however, all participants that filled out the survey were included in the total seizure count.

## Results

The current study involves 58 participants; of these, 43 caregivers were interviewed using the Vineland-3 and answered a survey regarding therapy and other questions (see Methods), 10 of whom completed the Vineland-3 but did not answer the survey, and 5 participants who answered the survey but have not yet performed the Vineland-3 due to language constraints. Included in the natural history analysis were all 53 who had at least one Vineland-3 assessment performed. A demographic breakdown of the specific pathogenic variants of the individuals included and their genders is present in **Table 1**. The variants are shown in **Figure 1**. Information on the racial breakdown by pathogenic variant can be found in **Supplementary Table 1.**

**Figure 1.**
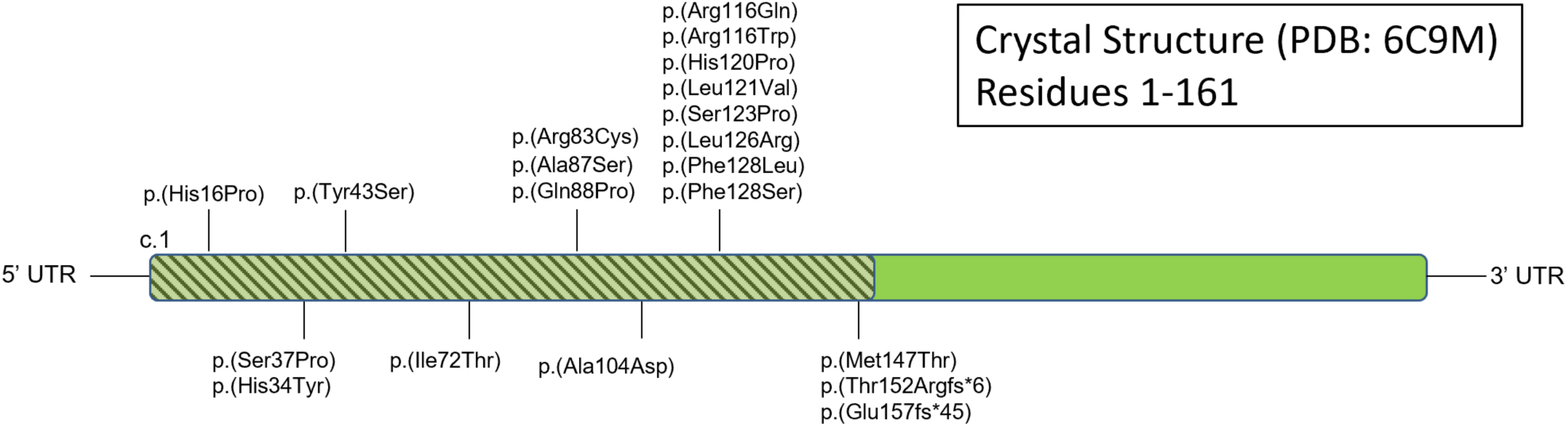
NAA10 missense or frameshiw variants from the cohort in this paper.

**Table 1.** Pathogenic variant breakdown by sex.

| Table 1. Pathogenic variant breakdown by sex. |  |  |  |
| --- | --- | --- | --- |
| Pathogenic variant | Females | Males | Total |
| His16Pro | 1 | 0 | 1 |
| His34Tyr | 0 | 1 | 1 |
| Ser37Pro | 0 | 1 | 1 |
| Tyr43Ser | 0 | 2 | 2 |
| Ile72Thr | 0 | 2 | 2 |
| Arg83Cys | 23 | 0 | 23 |
| Ala87Ser | 2 | 0 | 2 |
| Gln88Pro | 1 | 0 | 1 |
| Ala104Asp | 1 | 0 | 1 |
| Arg116Gln | 1 | 1 | 2 |
| Arg116Trp | 3 | 0 | 3 |
| His120Pro | 2 | 0 | 2 |
| Leu121Val | 1 | 0 | 1 |
| Ser123Pro | 1 | 0 | 1 |
| Leu126Arg | 1 | 0 | 1 |
| Phe128Leu | 7 | 0 | 7 |
| Phe128Ser | 1 | 0 | 1 |
| Met147Thr | 3 | 0 | 3 |
| Thr152Argfs*6 | 0 | 2 | 2 |
| Glu181Alafs*67 | 0 | 1 | 1 |
| Total | 48 | 10 | 58 |

The average age at time of most recent assessment was 12.4 years old with individuals ranging in age from 11 months to 40.2 years old. Across all major adaptive domains, there is a significant difference between individuals with *NAA10* pathogenic variants score lower and the normalized average Vineland score of 100 (σ=15) with Females performing more poorly than males. The average ABC and Domain standard scores for each Vineland Assessment recorded have been summarized by sex and pathogenic variant in Table 2. Growth Scale Value scores were also plotted against age for each Vineland subdomain and showed similar decrements in raw score in age as the standard scores (**Supplementary Figure 1**).

**Table 2.** Vineland ABC and Domain Standard Scores by Sex and Pathogenic mutation.

| Pathogenic variants | ABC | SD | Com | SD | DLS | SD | Socialization | SD | Motor | SD |
| --- | --- | --- | --- | --- | --- | --- | --- | --- | --- | --- |
| **Females (Average)** | **36.5** | **14.6** | **27.6** | **11.0** | **36.7** | **18.2** | **42.9** | **17.6** | **38.5** | **18.8** |
| p.His16Pro | 28.5 | 6.2 | 27.5 | 8.7 | 20.0 | 0.0 | 36.3 | 9.8 | * |  |
| p.Arg83Cys | 34.0 | 12.4 | 25.7 | 9.5 | 32.9 | 15.1 | 41.2 | 16.6 | 34.5 | 14.6 |
| p.Ala87Ser | 33.0 | 14.7 | 24.5 | 7.7 | 35.8 | 18.6 | 36.5 | 17.8 | 36.0 | 22.6 |
| p.Gln88Pro | 49.0 | --- | 32.0 | --- | 54.0 | --- | 58.0 | --- | 21.0 | --- |
| p.Ala104Asp | 45.0 | 11.3 | 26.7 | 9.9 | 46.3 | 17.8 | 59.0 | 13.1 | * |  |
| p.Arg116Trp | 44.8 | 18.2 | 32.0 | 14.2 | 44.3 | 28.1 | 55.5 | 12.1 | 68.0 | --- |
| p.His120Pro | 20.0 | 0.0 | 20.0 | 0.0 | 20.0 | 0.0 | 20.0 | 0.0 | * |  |
| p.Leu121Val | 52.0 | 0.0 | 45.0 | 7.1 | 43.0 | 5.7 | 65.0 | 1.4 | * |  |
| p.Ser123Pro | 57.7 | 3.8 | 42.3 | 11.7 | 61.3 | 1.2 | 65.7 | 4.0 | 68.7 | 4.0 |
| p.Leu126Arg | 66.0 | --- | 40.0 | --- | 94.0 | --- | 63.0 | --- | 49.0 | --- |
| p.Phe128Leu | 33.9 | 14.9 | 24.9 | 11.6 | 37.4 | 18.1 | 38.3 | 17.2 | 23.0 | 6.5 |
| p.Phe128Ser | 57.0 | 2.6 | 43.3 | 5.0 | 64.3 | 2.1 | 59.0 | 4.6 | 63.3 | 4.2 |
| p.Met147Thr | 35.1 | 18.5 | 28.9 | 15.1 | 35.9 | 18.2 | 38.9 | 22.1 | 57.0 | 0.0 |
| **Males (Average)** | **62.8** | **22.9** | **58.5** | **26.8** | **61.8** | **25.8** | **69.3** | **23.4** | **62.5** | **21.8** |
| p.His34Tyr | 76.0 | 1.4 | 75.5 | 0.7 | 80.5 | 10.6 | 78.0 | 2.8 | 77.5 | 12.0 |
| p.Ser37Pro | 40.0 | --- | 26.0 | --- | 40.0 | --- | 50.0 | --- | 20.0 | --- |
| p.Tyr43Ser | 32.8 | 4.1 | 31.0 | 2.6 | 26.5 | 7.5 | 38.5 | 3.9 | * |  |
| p.Ile72Thr | 80.0 | 18.5 | 76.7 | 28.3 | 76.3 | 14.2 | 91.3 | 21.4 | 58.5 | 2.1 |
| p.Arg116Gln | 58.0 | 7.1 | 40.0 | 5.7 | 62.0 | 11.3 | 68.5 | 6.4 | 59.0 | 12.7 |
| p.Thr152Argfs*6 | 63.0 | 8.5 | 54.0 | 8.5 | 70.5 | 16.3 | 62.5 | 2.1 | 53.0 | 21.2 |
| p.Glu181Alafs*67 | 87.3 | 12.7 | 91.7 | 7.4 | 83.3 | 24.9 | 94.0 | 6.0 | 85.5 | 20.5 |
| **Total Average** | **40.4** | **18.5** | **32.2** | **18.1** | **40.5** | **21.4** | **46.8** | **20.7** | **43.8** | **21.7** |

\* Motor scores were unable to be collected for individuals with the p.His16Pro, p.Tyr43Ser, p.Ala104Asp, and p.His120Pro pathogenic variants due to differences in the protocol at the time of collecting data.
--- Standard Deviation was unable to be calculated for pathogenic variants where n = 1.

In addition to the overall decreased behavioral development, individuals with an NAA10 pathogenic variant also showed a decrease in Adaptive Behavior over time. The evolution of ABC standard scores over time is seen in **Figure 2**. The overall trends show that as individuals with NAA10-related neurodevelopmental syndrome grow older, their function declines. This trend was shown in the most prevalent pathogenic variant, p.Arg83Cys (**Figure 2A**), in females with other pathogenic variants (**Figure 2B**), and in males (**Figure 2C**). These results are very similar when analyzed by Vineland subdomain scores (**Figure 3**), although the number of data points for males is low.

**Figure 2.**
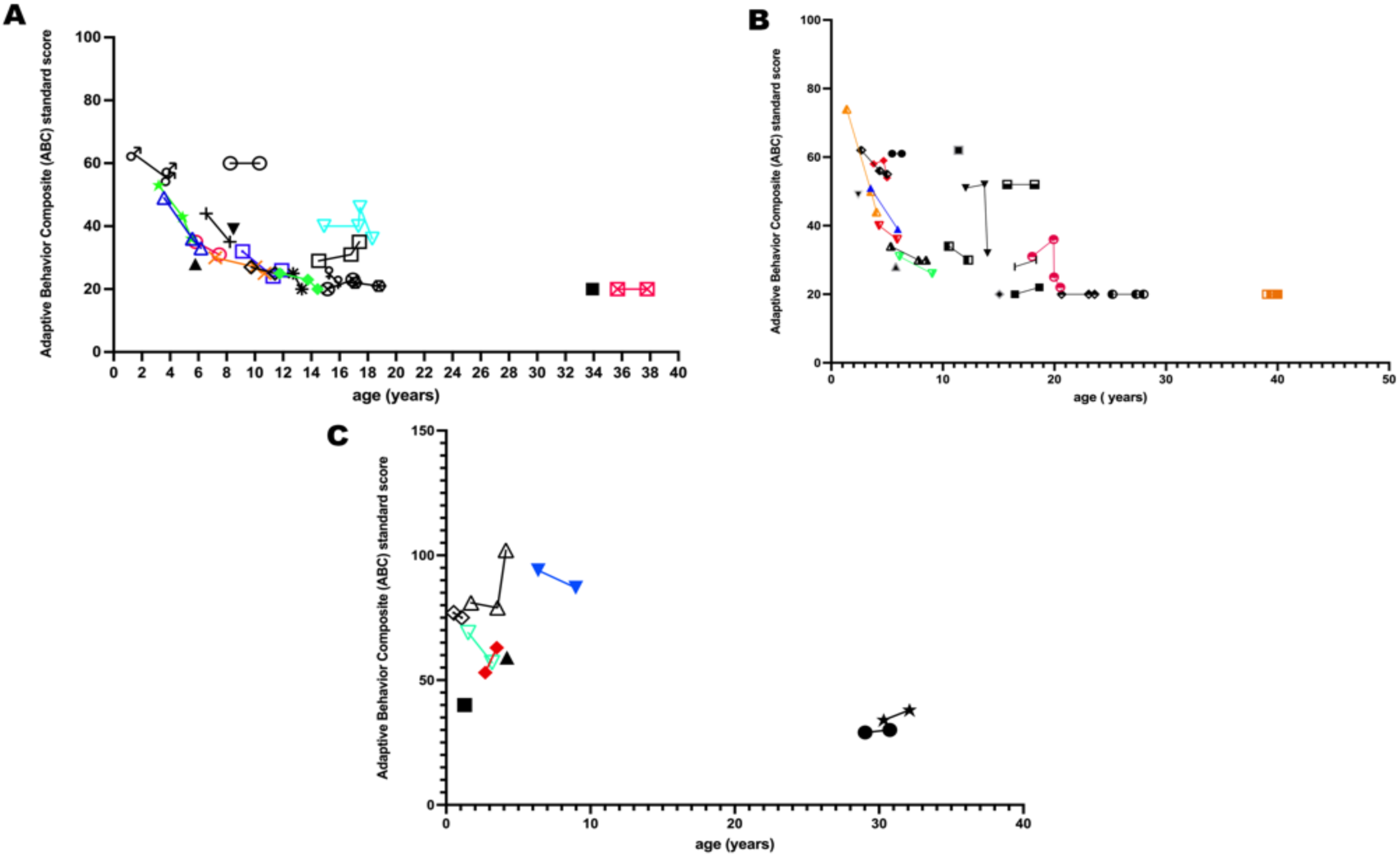
Participant’s ABC standard score as a function of their age at the time of assessment in years. Each icon represents one participant’s test taking session, the same icons connected by a line represents a participant’s scores over time. Figure A includes all female participants with the same mutation (Arg83Cys), Figure B is females with all other types of pathogenic variants and Figure C is all males with varied pathogenic variants. All 3 Figures show a decline in participant score over time, with some males being the exception.

**Figure 3.**
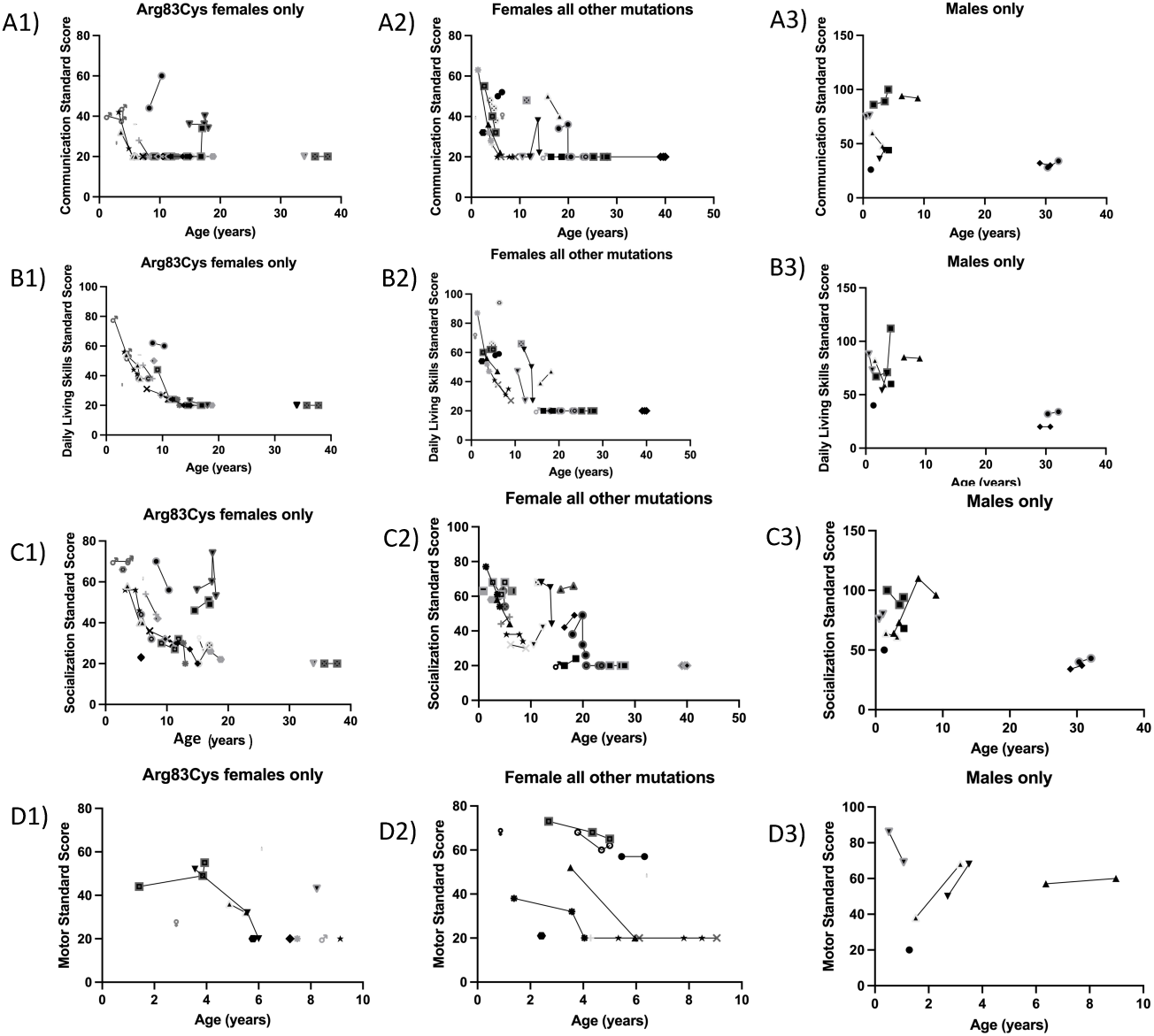
Individual Vineland Adaptive Domain scores were graphed over time. Each different type of dot is a different individual. Lines connecting dots show the evolution of a patients’ score after repeated Vineland assessments. Figures A1-3 showcase communication standard scores, B1-3 show daily living skills standard scores, C1-3 are socialization standard scores and Figures D1-3 are all available motor standard scores. All Figures are further filtered by gender and mutation. All Figures (1) include all female participants with the same mutation (Arg83Cys), Figures (2) showcase females with all other types of pathogenic variants and Figures (3) are males only with varied pathogenic variants. Visualization of each of the main adaptive domains was performed to determine if there was a particular area that had a greater detrimental effect to the overall development of these individuals and is shown. Communication, Daily Living Skills, and Socialization scores of Females with both p.Arg83Cys pathogenic variants and all other pathogenic variants follow a similar decrease in aptitude over time with age as did the ABC standard score. The Motor standard score does not appear to follow that trend. Although outliers display higher than average scores, their progress over time seems to follow the same downward trend as other participants.

Of the 48 patients that completed the survey, 9 had seizures. The specific pathogenic variants associated with the seizure phenotype can be seen in **Supplementary Figure 2** and **Supplementary Table 2**. Other qualitative information regarding the seizures types and the therapies used can also be found in **Supplementary Tables 5** and **6** respectively. The average age of individuals with seizures was 13 years old with a range from 4-39 (**Figure 4**). There was no significant difference (p = 0.3) in Vineland ABC Standard Score between the seizure group (μ = 35.6, σ = 14.0) and the non-seizure group (μ = 42.1, σ =19.0). Additionally, there was no significant difference found between the groups when comparing Vineland adaptive domain or sub-domain standard scores (**Supplementary Table 3**). There was no significant difference between when patients in the seizure and non-seizure groups achieved motor or language milestones (**Supplementary Table 4**). Information on the seizure types can be found in **Supplementary Table 5**. A summary of non-pharmaceutical and pharmaceutical therapies used by parents can be seen in **Supplementary Table 6**.

**Figure 4.**
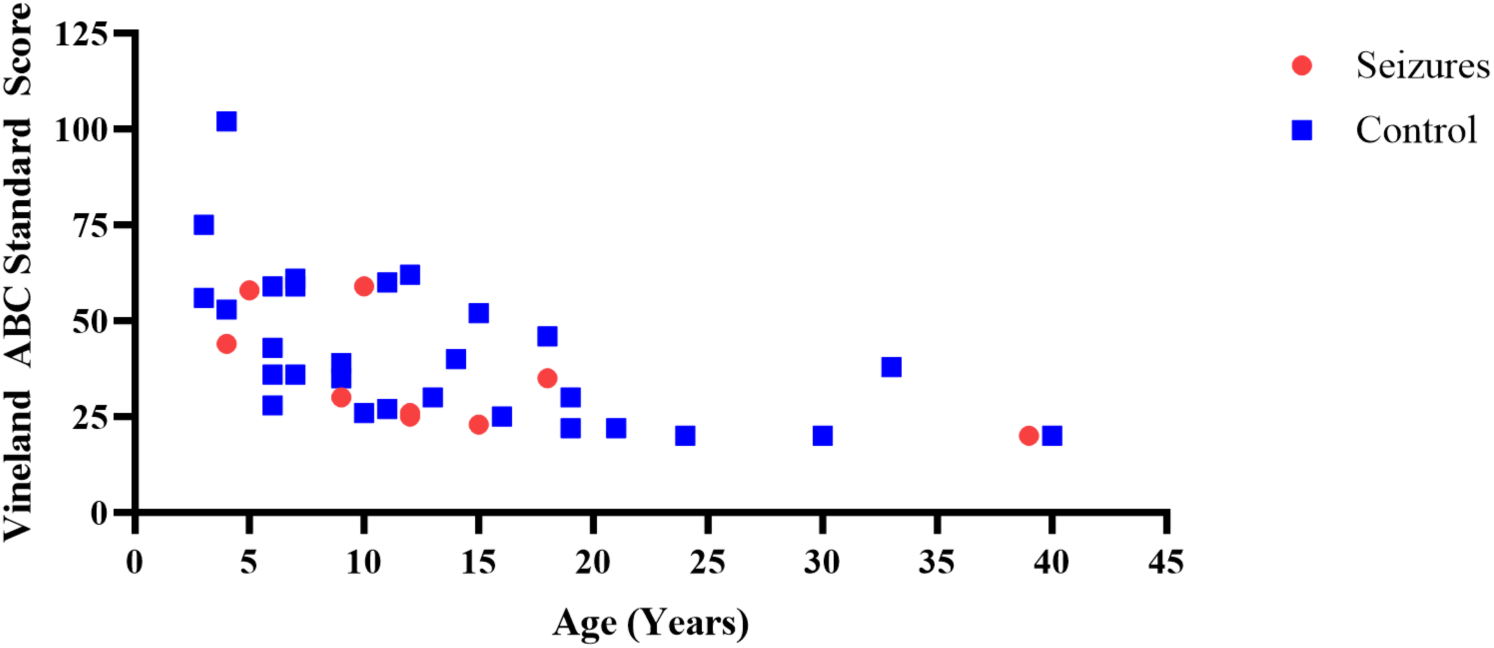
ABC Standard Score vs Age at time of Vineland Assessment based on Participant Seizure Status. The most recently administered Vineland ABC Standard scores vs the age at which the exam was administered for individuals with and without the seizure phenotype. Both individuals with and without seizures appear to be declining in function while following similar trajectories.

Information about types of therapy interventions used by participants was gathered from the survey (**Figure 5**). Speech therapy was the most widely used type, with a total of 39 participants, followed by physical therapy with 37 participants and occupational therapy with 32 participants. Less-used therapies include equine therapy with 17 participants, water therapy with 14 participants, ABA therapy with 9 participants and lastly, group therapy with 6 participants. A total of 17 participants reported using other non-pharmaceutical interventions, including art therapy, sensory integration therapy, chiropractors and others. A full list of therapies reported under ‘other’ can be found under **Supplementary Table 7**. Speech therapy had the most reports of ‘not helpful’, with 9 caregivers having expressed their dissatisfaction for various reasons. Other therapies only had 1-3 reports of ‘unhelpful’ by caregivers, with water therapy being the only therapy all caregivers reported being satisfied with. Comments from caregivers about why they thought therapies were helpful or unhelpful can be found under **Supplementary Table 8**.

**Figure 5.**
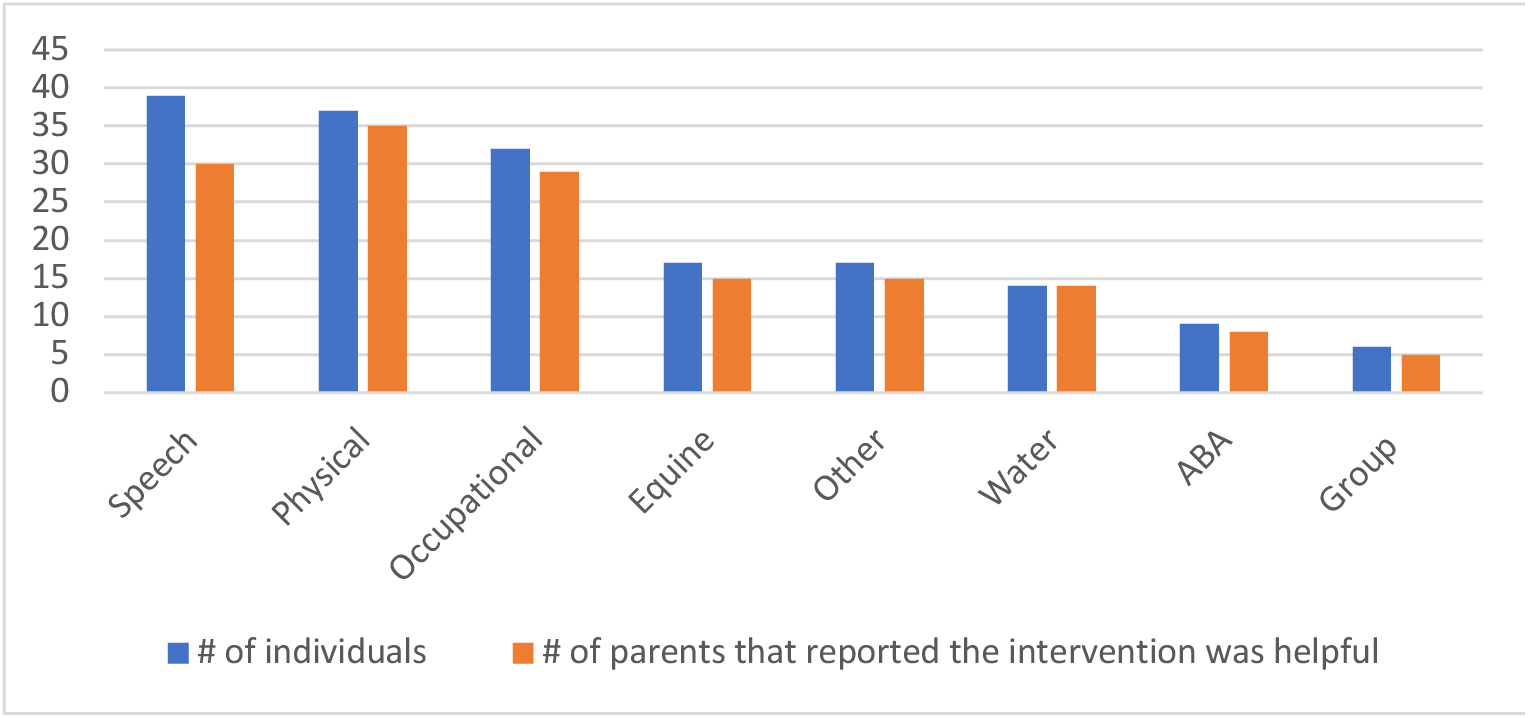
Therapy interventions used by individuals with NAA10-related neurodevelopmental syndrome and the number of parents that reported the intervention was helpful. Speech, physical and occupational therapy were the three most used types of therapy with over 30 participants in each, double the amount of other, less-used therapies. However, they were reported by caregivers to be less helpful than other therapies, with Speech therapy having the most reports of ‘not helpful’ followed by Occupational therapy. Water therapy was the only therapy that was reported by all participants to be helpful. Breakdown of therapies listed under ‘Other’ can be found in the supplementary information.

The number of therapies being received was counted for each individual, with 0 representing participants who are not receiving any type of therapy, and 8 being the most one can receive, including all 7 types of therapy defined previously plus ‘other’ if the participant received other non-pharmaceutical interventions. Please note that, while caregivers could include as many interventions as they wanted, they were all clumped into 1 type under ‘Other’ for the sake of organization, therefore multiple ‘other non-pharmaceutical interventions’ were not counted in this graph. Only the most recent ABC score of each participant was used. This analysis was done in order to identify whether receiving more or less types of therapy impacted participant’s ABC score or vice-versa (**Figure 6**). There was a significant correlation between ABC score and total number of therapies (p=0.0069), with higher scores being associated with lower amounts of therapy, and lower scores being associated with higher amounts of therapy, thus seeming to indicate that the lower-functioning individuals were being enrolled in more types of therapy.

**Figure 6.**
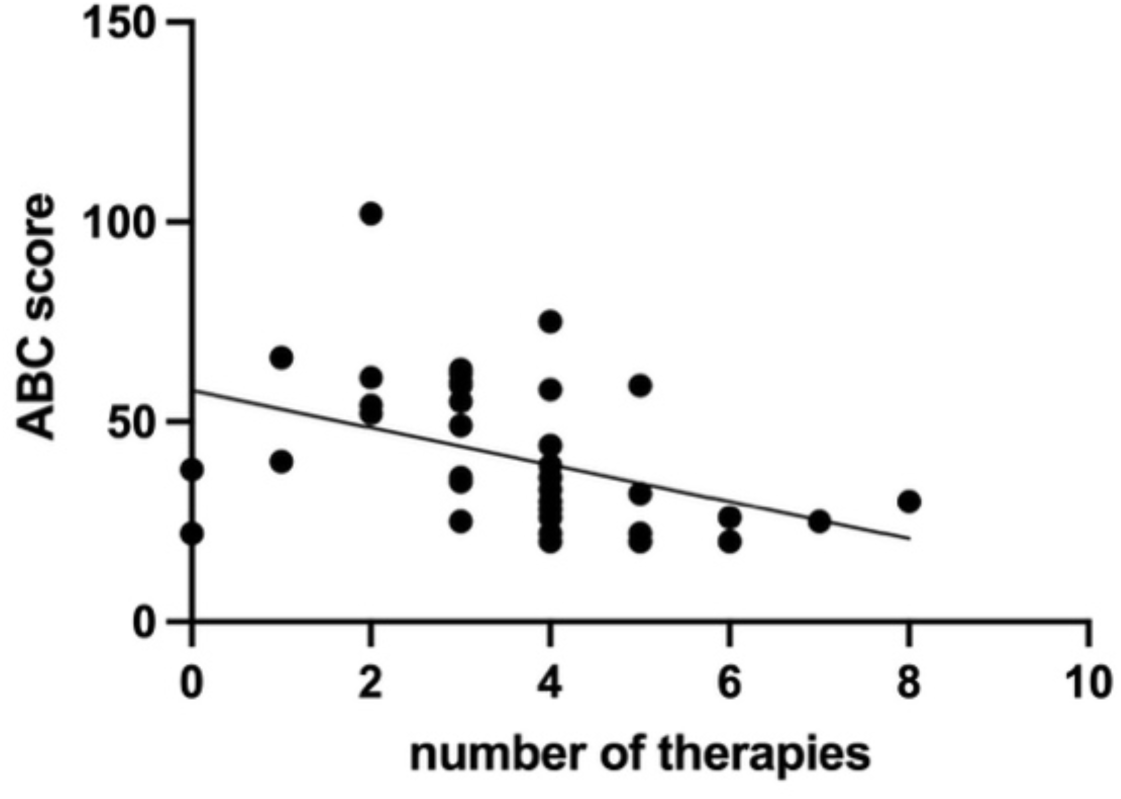
ABC scores vs. the total number of therapy interventions the individual receives. Only scores of their most recent test were used. The data shows significant correlation between scores and total amount of therapy (p=0.0069), where lower scores are associated with higher numbers of therapy. The data suggests more severely affected individuals receive more types of therapy.

Analysis between age at start of therapy and their most recent ABC score was done to determine whether earlier intervention had an impact on adaptive behavior (**Figure 7**). Age was represented in months in order to better include participants who had started therapy before the age of 1. Correlation analysis was only significant for speech therapy (p=0.0031) (**Figure 7A**), indicating that starting speech therapy at a younger age seems to help and might result in a better outcome. Correlation analysis of all other therapies was non-significant, with the following values: occupational therapy, p=0.68 (**Figure 7B**); physical therapy, p=0.76 (**Figure 7C**); ABA therapy, p=0.32 (**Figure 7D**); equine therapy, p=0.39 (**Figure 7E**); water therapy, p=0.14 (**Figure 7F**). Group therapy was not included due to a lack of data points. The data point for participant OS_118 (age range 40-44 years) was deleted from physical therapy for skewing the data extremely and making it difficult to see the pattern of other participants. Additional analysis was performed for each therapy using its corresponding sub-score, which did not find any significant correlations (**Supplementary Figure 3**). This was done in order to take a deeper look into early intervention and how it might impact its corresponding sub-domain of adaptive behavior. As speech therapy targets language and communication of all sorts, communication scores were used, rendering a p-value of 0.06 (**Supplementary Figure 3A**). Occupational therapy aims to help individuals learn, improve and maintain skills necessary to live independently, therefore daily living skills sub-scores were chosen for this therapy; analysis was non-significant with p=0.59 (**Supplementary Figure 3B**). Motor scores were used to assess physical, equine and water therapy due to its gross motor components, however the lack of data points makes it difficult to determine validity. Analysis of all three therapies were non-significant, with the following p-values: physical therapy, p=0.94 (**Supplementary Figure 3C**); equine therapy, p=0.98 (**Supplementary Figure 3D**). A p-value for water therapy could not be computed due to the lack of data points and variability between scores (**Supplementary Figure 3E**). Reasoning and further deliberation about the lack of motor scores can be found in the discussion section. Applied Behavior Analysis (ABA) encompasses an array of skills that is practiced in each session, therefore we analyzed multiple sub-domains within ABA, including communication, daily living skills and socialization (which can occur between the client and the behavior therapist, or in group settings if sessions occur in clinics). Analyses were all non-significant, with the following p-values: communication, p=0.59; daily living skills, p=0.29; socialization, p=0.18 (**Supplementary Figure 3F**).

**Figure 7.**
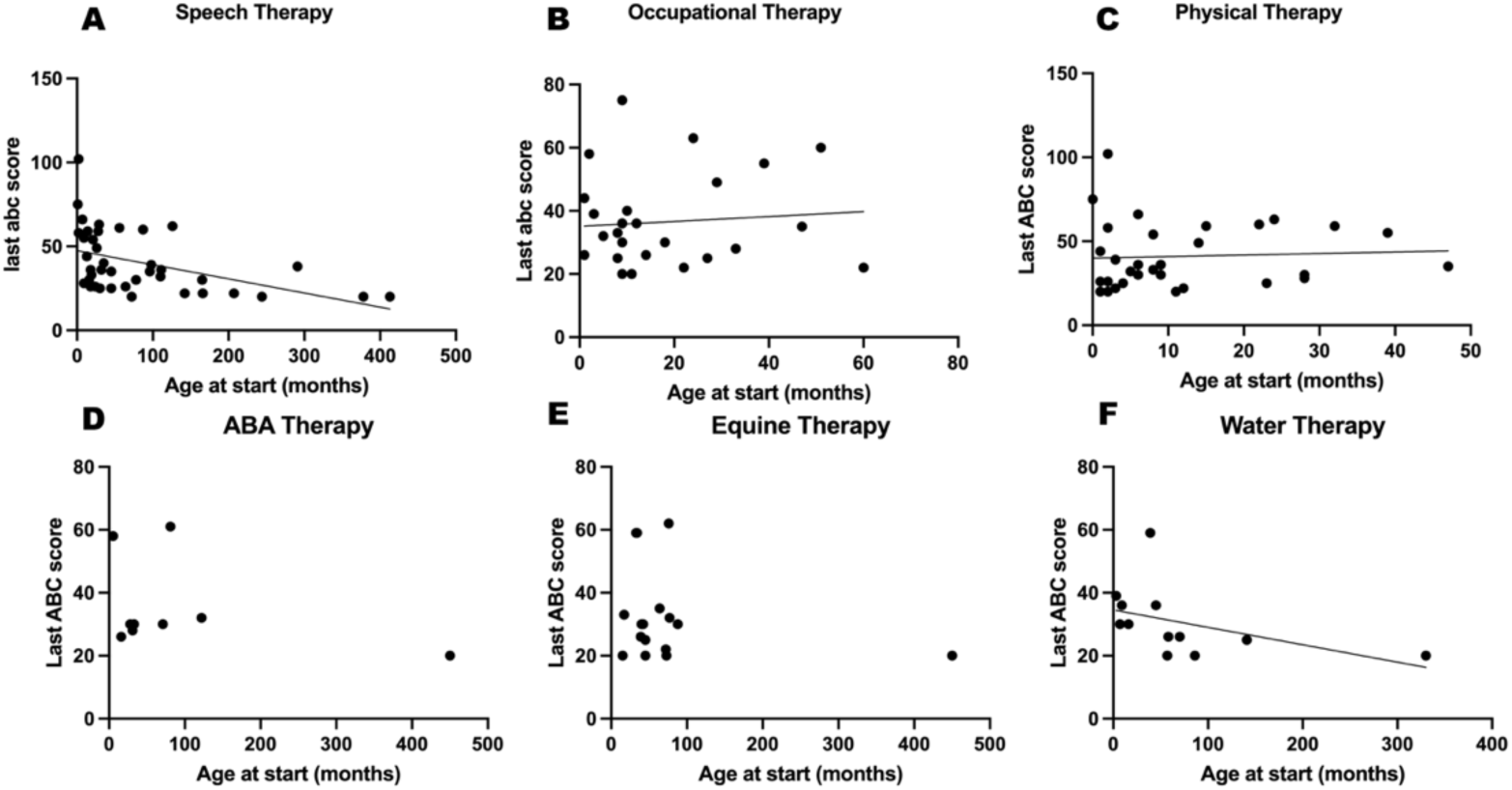
ABC scores as a function of the age the individual started the therapy (in months). Only scores of their most-recent test were used. Each Figure corresponds to the therapy indicated by the title. Correlation analysis of Figure A showed significance (p=0.0031), suggesting that starting speech therapy earlier in life may correspond to higher ABC scores. Correlation analysis of all other Figures was non-significant.

**Supplementary Table 7** consists of all therapies reported by caregivers under ‘other’, which gave them the space to cite any other non-pharmaceutical interventions their child may have received at any time. More commonly used therapies, such as visiting a chiropractor (3 participants), were separated into its own category with multiple participants for better visualization. Less used therapies were only separated if the caregiver made specific comments about each. Otherwise, all therapies reported by the same caregiver were included together as some caregivers made general comments. All information was de-identified, names previously cited have been replaced by [NAME] or the appropriate pronouns. The wording in comments was only changed if there were grammar and language errors to be corrected. **Supplementary Table 8** includes comments from caregivers about why they thought certain therapies were helpful/unhelpful. Comments were chosen without specific criteria, other than being descriptive as opposed to comments with little to no description.

## Discussion

Individuals with Ogden syndrome performed significantly worse than the mean standardized score on the Vineland 3 Assessment. When performing analysis by sex, females performed more poorly than males. Due to the X-linked inheritance of this disease, it stands to reason that males should present with a more severe phenotype, regardless of genotype^15,28^. However, even when comparing individual pathogenic variants present in the few surviving males, they score as well or better than the females on average. A possible explanation for this could be that males that survive infancy have a less pathogenic variant, with some of these males inheriting a mutation from a female carrier who are themselves either unaffected or minimally affected, due to that particular variant being overall much less deleterious to protein function. Possible evidence supporting this explanation can be seen when comparing the Vineland Scores between males with the p.Tyr43Ser pathogenic variant and those with the p.Glu181Alafs*67 pathogenic variant. Amino Acid 181 is much closer to the 3’-UTR of the NAA10 exon than 43, allowing for more protein to be translated, with presumably some expression of the intact acetyltransferase domain (although this has not been formally tested, as there are no available cell lines yet established from these males). The construct used for the crystallization of NAA10 included residues 1-161 (**Figure 1**), and the remaining C-terminus is an unstructured region that has not been as thoroughly studied. It is possible that the C-terminus might undergo proteolysis/clipping to yield an intact, unaffected core enzymatic domain, but this needs further study. Additionally, while increased functional protein size may be a factor in the less pathological effect of this variant, it should be noted that males with the p.His34Tyr pathogenic variant also scored better than p.Tyr43Ser males suggesting that the nature of the missense change likely has an effect on enzymatic activity, either via altering expression level, stability, catalytic function, or ability to form the NatA complex, as was demonstrated in prior publications^15,37,50^. Conversely, females might survive more deleterious pathogenic variants than the males, due to being heterozygous with a fully functioning allele and also subject to skewed X-inactivation^15,21^. Furthermore, the sample of males available for testing (N=17) was much smaller than that of the females (N=97). Given both groups showed similar declines in function as they increased in age, it is possible that the males will exhibit similar Vineland Scores when compared at similar ages to their female counterparts as more data points are collected in the future.

In order to determine if there was a specific adaptive domain that individuals with NAA10-related neurodevelopmental syndrome were especially deficient in, Vineland scores were separated by the Adaptive Domains. Once again, on average, individuals with Ogden syndrome performed poorly compared to the normalized Vineland Domain Standard Scores. The earlier difference present between the sexes also remained. One thing of note that was different between the four adaptive domains was that the Motor Domain score did not seem to have the same decline as the other domain scores. This suggests that the motor skills of individuals with NAA10-related neurodevelopmental syndrome may not be as adversely affected as their Communication, Socialization, and Activities of Daily Living skills. This stands in contrast to some of the current phenotypic descriptions of this disease with some individuals having low muscle tone and difficulty walking^15,24,37^. It seems to be more likely that the abnormal motor scores collected are due to variation in how the assessments were originally collected. Pearson suggests that the Motor Skills Domain of the Vineland 3 Assessment is normative for those ages 0-9^49^. However, this range is based off the disorders that the assessment is rated for. Given the rarity and severity of Ogden syndrome, it was decided to collect Motor Domain information regardless of the test taker’s age. As more Motor Domain information is collected, it is possible that the difference in magnitude between its scores and the other adaptive domain scores will decrease.

The Communication Adaptive Domain standard score in the p.Arg83Cys females also matches the decline exhibited by the ABC standard score over time. However, there is an outlier present that seems to improve in communication score from their first (com ss = 44) to second (com ss = 60) Vineland administration. They also perform significantly better than their peers at both time points. However, this individual did not see a analogous increase in their ABC score due to their Socialization Domain score dropping from 70 to 56 in the same two year time frame between when the assessment were administered. While this was an overall decrease in their Socialization standard score, they still scored better than their similarly aged peers at both time points. Given the patient was between the age ranges of 6-10 years old when given the assessments, it will be interesting to see how their scores change in the future. If they stagnate or improve, further interviews with the patient and their caregivers may be warranted to determine what extraneous factors, if any, contributed to their increased function compared to their peers.

There were no significant differences found between the different genotypes for Ogden syndrome and having the seizure phenotype. However, due to the small sample size, it is too soon to determine if there is zero association between seizures and the different genotypes for Ogden syndrome. However, despite the lack of significance, the population of patients with seizures matches the current population incidence of seizures^15^. Across the Vineland Standard Score broad and sub-domains, there was no significant difference between the seizure group and the non-seizure groups. This could suggest that seizures are not associated with developmental delays in NAA10-related neurodevelopmental syndrome. However, this is unlikely due to the breadth of literature that suggests that the incidence of general developmental delay and seizure are positively correlated^44–47^. It is a possibility that the rapid treatment intervention received by the participants was sufficient in decreasing these increased likelihoods in developmental delay as there is an association between the time of diagnosis of epilepsy and decreased Vineland scores^44^. However, it is more likely that increasing the sample would decrease the variability within the results allowing for discrepancies in the data to be seen. There was no significant difference between when the individuals in the seizure group and the non-seizure group achieved various developmental milestones.

The trend in ABC standard scores in participants with seizures matched the overall trend found in the natural history analysis performed. As individuals with Ogden syndrome age, they tend to score more poorly on Vineland Assessments. Even patients whose caregivers discussed having their child go into remission after treatment of their epilepsy did not present as outliers on the graph. However, the analysis performed was limited by a few factors. Having a sample size of only 44 eligible participants made between-participant analysis highly variable. Further experimentation should aim to increase the number of participants. Additionally, of the participants who had seizures, there was no uniformity in the types of seizures they had. This detracts from the generalizability of the seizure group data making conclusions less likely to be valid. Continued experiments should aim to rectify this issue by grouping individuals by the type of seizure that they had and cross-validating the results with the overall seizure group. This type of analysis was not performed in this experiment due to uncertainty surrounding the type of seizure that the participant had, thus highlighting a need to collect much more data specifically about the seizures in the future. Additionally, for the participants that had a diagnosis for their seizure type, there were not enough of them within a group to make comparisons. There is also a lack of longitudinal Vineland data for this experiment meaning that comparing the longitudinal adaptive behavioral scores of the participants over time was not possible as many of them had only received a baseline and secondary Vineland assessments. With greater amount of longitudinal data, more robust comparisons can be made to determine if there are differences in developmental timelines between the non-seizure group and seizure groups. Additional areas of focus that should be investigated further in the next iteration of this study could be to query caregivers on the length of time between when they noticed the child had their first seizure and the child started preventative treatment for them given the association between decreased Vineland scores and an increased time to diagnosis of epilepsy^44^.

The general picture of the types of therapy interventions being used by individuals with NAA10-related neurodevelopmental syndrome is shown in **Figure 5**, and caregivers had the option to report the therapy as helpful or unhelpful. Speech, occupational and physical therapy, respectively, were the most widely used types of therapy within our cohort, which exactly corresponds to other previously published data on individuals with ASD^51^. Other therapies, such as equine, ABA and water therapy were less used, but caregivers reported being mostly satisfied with them which could encourage other caregivers to try these therapies. Previous studies have reported beneficial effects of equine therapy on behavior and social communication in individuals with ASD who participated in the therapy^52^ as well as gross motor skills^53^. Similarly, water therapy has been shown to improve motor skills of persons with disability^54,55^ as well as emotional response, adaptation to change and overall improvement of functional impairments seen in ASD^56^. ABA has also been previously linked to improvements in adaptive behavior in individuals with ASD^57^.

As seen in **Figure 6**, higher-functioning participants receive less therapy than lower-functioning individuals, suggesting more severely affected individuals and their caregivers are looking for more alternative treatments in addition to standard treatments. A similar result has been shown in individuals with ASD, where alternative types of care are mostly used by severely affected individuals and individuals with additional problems^58^. This relates to the concern that mainstream care for people with severe disabilities, including Ogden syndrome, is deficient in certain aspects.

Early intervention refers to starting a therapy treatment as early in life as possible. Early intervention has been linked to significant improvements in children with neurodevelopmental disorders^59^. The myriad reasons for its effectiveness are outside the scope of this paper, but comprehensive descriptions have been previously published^60^. In our study, early intervention was only significant for speech therapy, suggesting starting speech therapy earlier in life may lead to better outcomes. However, the lack of significance for other therapies could be due to smaller sample sizes. Speech therapy was, in fact, the therapy in our study with the greatest number of participants. Other therapies had as little as 8 participants, as was the case for ABA, and group therapy was not included in the analysis due to the lack of data points. Sub-score analysis was non-significant across all therapies. However, the p-value for the correlation between age at start of speech therapy and communication standard scores was very close to significance (p=0.06). Additional data could possibly bring this value closer to significance, so it is important to revisit this analysis in the future with more participants. The lack of motor scores also made a huge impact in the analyses where motor components were the most important. Equine and water therapies were added for record-keeping purposes, but also highlight the need for a more in-depth analysis later on. Future research in the field should continue to investigate early intervention across all therapies, aiming to gather as many participants as possible.

Overall, there is not enough data to suggest therapies are helping individuals improve. The adaptive behavior is still declining over time despite the therapies being received. ABC scores were used throughout analyses for the sake of standardization, and are a good measure of adaptive behavior overall. However, because ABC scores are norm-referenced, they fail to highlight any improvement made on an individual level. Growth-scale value (GSV) scores are non norm-referenced and therefore might highlight individual improvements over time^61^. A preliminary analysis of GSV scores of individuals in our cohort with the pathogenic variant Arg83Cys can be found in the supplementary information. Although analyzing GSV scores instead of ABC scores can help with the floor effect seen in ABC analyses, it does not completely eliminate it. GSV score analysis also fails to give a comprehensive measurement, as it is divided into 11 sub-categories. Future research can aim to comprehensively investigate the relationship between therapies and GSV scores in order to understand the improvements seen at the individual level and how much of such improvements can be attributed to the therapies being received. Unfortunately, for most participants, we did not have ABC scores from before they started therapy to compare before-and-after results. Ideally, a prospective study that follows participants throughout their therapy interventions could better showcase the improvements made in that period of time, although it is very difficult to fund such studies long-term, particularly for ultra-rare genetic disorders like this one.

Given current therapeutic interventions aimed at improving adaptive behavior used by caregivers of individuals with Ogden syndrome are largely ineffective, alternative treatments should be considered. Current pharmaceutical treatments that have been beneficial in improving function in autism spectrum disorder and Fragile X Syndrome should be considered as possibilities to manage symptoms or improve outcomes. Serotonergic medications, specifically selective serotonin reuptake inhibitors have been shown to increase cognitive, expressive language, and motor function in individuals with both autism spectrum disorder and Fragile X Syndrome^62–65^. Additional research should also begin to focus on molecular and gene therapy to rescue protein function. Gene therapy has shown promising results in restoring function in mouse models of disease. Currently, adeno-associated viral vectors carrying plasmids with the gene of interest are the most common way of restoring protein functions in-vivo^66^. With the advent of CRISPR-Cas9 technology and other strategies for “knock-in” genome editing^67–69^ as well as advancements in RNA editing^70^, antisense oligonucleotide gene rescue^71^, X-chromosome reactivation^72^, and therapeutic nanoparticles^73^ there will be further avenues to explore in restoring NAA10 function. Despite the difficulties involved in developing a vector able to cross the blood brain barrier can pose a challenge^74–77^, there are currently therapies being developed for other neurodevelopmental disorders. For example, gene therapies for mouse models and cell lines in Fragile X Syndrome^78^, Rett Syndrome^79^, Angelman Syndrome^80^, and others^81–84^ have all completely or partially restored protein function. These results suggest that a restoration of function and improved adaptive behavioral outcomes over time are possible if implemented early enough.

## Conclusion

Ogden syndrome is a constellation of symptoms that encompasses anatomical defects, physiological dysfunction, and severe intellectual and behavioral delays. The severity across genotypes tends to initially present itself as sex-dependent where, paradoxically, females with the disease are having greater struggles than the few surviving males of the same age. However, this difference in development drops off with age as both groups appear to achieve similar Adaptive Behavioral Development scores over time, with the caveat that some of the C-terminal truncating variants in males may have overall better functioning due to retaining possibly an intact acetyltransferase enzyme domain. Despite the breadth of therapies that current caregivers have been trying to slow or reverse the chronology of symptoms, there seems to be no effective treatment yet found. Further research must be done with collecting more longitudinal Vineland behavioral data and expanding the overall cohort numbers.

## Data Availability

All data produced in the present study are available upon reasonable request to the authors

## Author Contributions

GJL and RH conducted all virtual interviews with participants and were responsible for primary Vineland data collection, with data curation conducted by EM. RM and CC were responsible for collection of survey data, data analysis, and project conception, along with GJL. The first draft of the manuscript was written by RM and CC, with critical revision performed by GJL at several points.

## Acknowledgments

We thank the families and the foundation, Ogden CARES for their participation and support. Leah Gottlieb provided assistance with Figure 1. Several medical students, including Andrew Trandai and Travis Beisheim, assisted EM with data curation.

## Ethical Approval

Both oral and written patient consent were obtained for research and publication, with approval of protocol #7659 for the Jervis Clinic by the New York State Psychiatric Institute - Columbia University Department of Psychiatry Institutional Review Board.

## Funding

This work is supported by New York State Office for People with Developmental Disabilities (OPWDD) and NIH NIGMS R35-GM-133408.

## Competing Interests

The authors declare that they have no competing interests or personal relationships that could have influenced the work reported in this paper.

## Notes

### Competing Interest Statement

The authors have declared no competing interest.

