## Supplementary Information for "Longitudinal Adaptive Behavioral Outcomes in Ogden Syndrome by Seizure Status and Therapeutic Intervention"

**Supplementary Table 1. Mutation breakdown by participant race and ethnicity**

| Mutation | Caucasian | Black | Asian | Hispanic/Latino | Total |
|---|---|---|---|---|---|
| p.His16Pro | 1 | 0 | 0 | 0 | 1 |
| p.His34Tyr | 0 | 1 | 0 | 0 | 1 |
| p.Ser37Pro | 1 | 0 | 0 | 0 | 1 |
| p.Tyr43Ser | 2 | 0 | 0 | 0 | 2 |
| p.Ile72Thr | 2 | 0 | 0 | 0 | 2 |
| p.Arg83Cys | 18 | 1* | 4 | 1 | 23 |
| p.Ala87Ser | 2 | 0 | 0 | 0 | 2 |
| p.Gln88Pro | 1 | 0 | 0 | 0 | 1 |
| p.Ala104Asp | 1 | 0 | 0 | 0 | 1 |
| p.Arg116Gln | 2 | 0 | 0 | 0 | 2 |
| p.Arg116Trp | 3 | 0 | 0 | 2 | 3 |
| p.His120Pro | 2 | 0 | 0 | 0 | 2 |
| p.Leu121Val | 1 | 0 | 0 | 0 | 1 |
| p.Ser123Pro | 1 | 0 | 0 | 0 | 1 |
| p.Leu126Arg | 1 | 0 | 0 | 0 | 1 |
| p.Phe128Leu | 7 | 0 | 0 | 0 | 7 |
| p.Phe128Ser | 1 | 0 | 0 | 0 | 1 |
| p.Met147Thr | 2 | 0 | 1 | 0 | 3 |
| p.Thr152Argfs*6 | 2 | 0 | 0 | 0 | 2 |
| p.Glu181Alafs*67 | 1 | 0 | 0 | 1 | 1 |
| Total | 51 | 2 | 5 | 4 | 58 |

**Supplementary Table 2. Mutations Associated with Seizures**

| Mutations | No Seizure | Seizure | Total (#) |
|---|---|---|---|
| p.His16Pro | 1 | 0 | 1 |
| p.His34Tyr | 1 | 0 | 1 |
| p.Ser37Pro | 1 | 0 | 1 |
| p.Tyr43Ser | 1 | 0 | 1 |
| p.Ile72Thr | 0 | 1 | 1 |
| p.Arg83Cys | 9 | 6 | 15 |
| p.Ala87Ser | 1 | 0 | 1 |
| p.Ala104Asp | 1 | 0 | 1 |
| p.Arg116Gln | 1 | 0 | 1 |
| p.Arg116Trp | 3 | 0 | 3 |
| p.His120Pro | 1 | 0 | 1 |
| p.Ser123Pro | 1 | 0 | 1 |
| p.Phe128Leu | 3 | 2 | 5 |
| p.Phe128Ser | 1 | 0 | 1 |
| p.Met147Thr | 3 | 0 | 3 |
| p.Glu181Alafs*67 | 1 | 0 | 1 |
| **Total** | **29** | **9** | **38** |

A numerical count of the different genotypes expressed by the individuals included in the survey.

**Supplemental Table 3. Seizure vs No Seizure for Vineland Assessment Standard Scores across all domains and subdomains**

| Seizures | | Age (years) | Seizure ABC SS | vi3_rec_ss | vi3_exp_ss | vi3_wrn_ss | vi3_per_ss | vi3_dom_ss | vi3_cmm_ss | vi3_ipr_ss | vi3_pla_ss |
|---|---|---|---|---|---|---|---|---|---|---|---|
| | Average | 12.6 | 35.6 | 3.2 | 1.3 | 2.6 | 1.0 | 3.8 | 2.9 | 3.9 | 3.1 |
| | STD | 10.0 | 14.0 | 3.3 | 0.9 | 2.3 | 0.0 | 2.7 | 2.6 | 3.4 | 3.1 |
| No Seizures | Average: | 12.1 | 42.1 | 4.2 | 2.9 | 2.5 | 3.1 | 5.6 | 3.6 | 6.0 | 4.3 |
| | STD: | 9.5 | 19.0 | 4.2 | 4.2 | 3.2 | 4.3 | 4.3 | 3.2 | 4.2 | 3.8 |
| | Welch's T-test | 0.9 | 0.3 | 0.5 | 0.1 | 1.0 | 0.0 | 0.2 | 0.5 | 0.2 | 0.4 |
| | Difference | 0.5 | -6.6 | -1.0 | -1.5 | 0.0 | -2.1 | -1.8 | -0.7 | -2.1 | -1.2 |

| Seizures | | vi3_cop_ss | vi3_gmo_ss | vi3_fmo_ss | vi3_int_ss | vi3_ext_ss | vi3_com_ss | vi3_dls_ss | vi3_soc_ss | vi3_mot_ss |
|---|---|---|---|---|---|---|---|---|---|---|
| | Average | 6.9 | 2.7 | 2.3 | 19.0 | 16.3 | 27.8 | 34.0 | 42.8 | 31.7 |
| | STD | 2.4 | 2.4 | 1.9 | 2.1 | 1.6 | 9.8 | 15.3 | 17.4 | 16.5 |
| No Seizures | Average: | 7.3 | 4.8 | 5.1 | 19.3 | 17.3 | 33.0 | 42.9 | 48.2 | 45.5 |
| | STD: | 2.8 | 3.9 | 4.7 | 1.4 | 2.6 | 19.1 | 21.8 | 19.4 | 23.7 |
| | Welch's T-test | 0.7 | 0.3 | 0.2 | 0.7 | 0.3 | 0.3 | 0.2 | 0.5 | 0.4 |
| | Difference | -0.4 | -2.1 | -2.7 | -0.3 | -1.0 | -5.3 | -8.9 | -5.4 | -13.9 |

The average recorded Vineland standard, domain, and sub-domain scales as compared between seizures and non-seizure groups. There was no significant differences between the Average scores between either group ($p < .05$).

**Supplemental Table 4. Developmental Milestones Achieved**

| **Seizures** | | At what age (in months) did you child first begin sitting on their own? | At what age (in months) did your child first begin crawling? | At what age (in months) did your child first begin walking? | At what age (in months) did your child speak their first word? |
|---|---|---|---|---|---|
| | Average | 13.4 | 39.0 | 33.0 | 28.5 |
| | STD | 5.1 | 25.2 | 13.1 | 14.3 |
| **No Seizures** | Average: | 19.5 | 20.2 | 33.4 | 21.6 |
| | STD: | 14.3 | 9.3 | 16.3 | 10.2 |
| | Welch's T-test | 0.1 | 0.2 | 1.0 | 0.5 |
| | Difference | -6.1 | 18.8 | -0.4 | 6.9 |

The average age at which developmental milestones were achieved between the seizure and non-seizure groups. There was no significant difference between the groups achieving their milestones ($p <.05$).

**Supplementary Table 5. Types of Seizure Reported**

| Seizure Type | Number |
|---|---|
| Absence | 2 |
| Focal | 3 |
| Drop | 1 |
| Myoclonic | 1 |
| Generalized Tonic Clonic | 3 |
| West Syndrome | 1 |
| Unsure | 5 |
| Total | 16 |

A numerical count of the types of seizures that participants recorded that their child had. A majority of patients were unsure of the specific type of seizure their child had. There does not appear to be a trend in focality or generalization of in the presentation of seizure post diagnosis. The total number of seizures is greater than the total number of participants due to some participants experiencing multiple types of seizure.

**Supplementary Table 6. Summary of Seizure Treatments**

| Initials | Current | Failed | Notes: |
|---|---|---|---|
| OS_115 | NR | NR | |
| OS_126 | NR | NR | |
| OS_112 | Onfi | Topamax, Lamictil, Epidiolex, Keppra, Valproic Acid, Phenytoin, Dilantin, Fintepla, high dose steroids & ketogenic diet. | Corpus callosotomy provided partial relief |
| OS_118 | Lamotrigine | None | |
| OS_119 | None | None | |
| OS_113 | Keppra, Epidiolex, depakote, CBD/thc | Clobazam | |
| OS_110 | NR | NR | |
| OS_151 | NR | NR | |
| OS_152 | Remission; Sabril | Keppra | Took Sabril per protocol for ~9 month and was able to stop her meds |
| OS_149 | Remission; Keppra | None | |

A summary table of the various treatments that each participant used to control their children's seizures. NR stands for not reported if a patient chose not to report or could not remember the types of medication their child was treated with. Patient response to drug types was variable but patients were able to enter a refractory period after finding the regimen that worked in their case.  The OS identification numbers, with the key to identify particular research participants, are only known to the study investigators.

| Supplementary Table 7. Parental Remarks on Therapies | | |
|---|---|---|
| **Type** | **OS number** | **Caregiver comments** |
| Respiratory physiotherapy | OS_160 | Helps [him] move his muscles and the mucus from his lungs to be able to breathe better and without [additional] oxygen. |
| Red light therapy | OS_151 | The red light therapy/intensives were beneficial at the time and [she] certainly seemed stimulated and happy but it was hard to maintain that level of care […] so it was not sustainable to continue to see [the therapist] consistently. |
| Naturopathic/holistic medicine | OS_151 | The holistic doctor/nutraeval was able to give us information on specific deficiencies [she] had so we could make dietary changes which was valuable. This doctors regimen most effectively controlled reflux long term for [her]. |
| Fish oil, CBD oil | OS_156 | Nothing really helped. I believe in fish oil and vitamins. I would probably believe massage could help her but there's no way [she] would stay still for it. |
| Chiropractor | OS_110 [1]<br>OS_113 [2]<br>OS_170 [3] | [1] Helped him learn how to walk by again. [2] Helps with overall health. [3] Tone, torticollis, coordination, movement in neck. Improved digestion. |
| Music therapy | OS_115 [1]<br>OS_112 | [1] Tried just one session but she didn't enjoy as she is noise sensitive. |
| Individual therapy with counselor or psychologist | OS_103 [1]<br>OS_169 [2] | [1] Her therapist talks to her about managing her stress and anxiety and what to do in social situations. [2] Helps my child's behavior. |
| Sensory Integration therapy | OS_162 [1]<br>OS_115 [2] | [1] Those practices intervened with [her] senses, for example she learned to smell things and she started to feel her feet and she got help seeing. [2] Works on her processing, regulation, developing vestibular and proprioceptive input, balance, rotation and she has a sensory diet in place throughout the day to help her have more awareness of her body and it helps to calm her. She is highly noise sensitive and visual processing is poor too, this therapy is the most important for help as it helps her to cope and function with all the other therapies and with the world around her better. |
| Adaptive dance/ice skating | OS_112 | These were great [stopped for personal reasons]. |
| Doman-Delacato method | OS_118 | [NAME] enjoyed the social aspect of the exercises, she did develop more physically and speech wise she did attempt the beginnings of names of helpers otherwise nothing greatly |
| Osteopathy, chromotherapy, tomatistherapy | OS_171 | With osteopathy we have corrected postural problems with the rest we have not noticed any positive change. |
| Neuroblend therapy (Roode draak Biddinghuizen), molli suit | OS_149 | She became stronger. |
| Syndromes without a name, swimming lessons, art therapy | OS_119 | N/a |

Types of non-pharmaceutical interventions used by individuals with Ogden Syndrome as reported by caregivers under the 'Other' section, and related comments.The numbers in [ ] next to the OS numbers corresponds to their respective comments about each therapy. The OS identification numbers, with the key to identify particular research participants, are only known to the study investigators

### Supplementary table 8. Comments from caregivers on why they believe therapies were helpful or unhelpful.

| Type | OS number | Helpful? | Comments |
|---|---|---|---|
| Speech | OS_143 | Yes | Her language skills have gradually improved and even now we see improvements. |
| Speech | OS_163 | Yes | Her spoken language is very limited and her comprehension significantly better. She needed to learn alternative ways to communicate. By age ten [she] was finally introduced to a speech generating device after demonstrating with computer games that her receptive language was so much better and that she responded to the voice output. After that it was important, and remains important to maintain and/or improve her skills in communicating however she can and using the available technology to assist her. |
| Speech | OS_153 | Yes | It helped her to make choices using items/pictures, to use a step-by-step communicator button to play recorded messages and to help with feeding/hand over hand feeding techniques. |
| Speech | OS_151 | Yes | At this point we are training on an eye gaze compatible assistive speech device which she is gaining some independence on and enjoying to communicate her needs. |
| Speech | OS_156 | Yes | Speech Therapy in general works on so many aspects of the person - social, cognitive/comprehension, and obviously communication. She uses AAC to speak but also likes to try to use her mouth so PROMPT therapy has been necessary for that. |
| Speech | OS_103 | Yes | Helped with feeding and swallowing when younger and communication skills now. |
| Speech | OS_148 | No | Lack of knowledge around diagnosis, lack of knowledge for AAC training. AAC was not supported, so we funded it ourselves and trained using online resources. |
| Speech | OS_162 | No | Vision challenges made it difficult to recognize photos which is the base level of communication and fine motor skills made it impossible to manipulate aac devices. Even when using switch devices there was no clear indication of understanding choices, more understanding we wanted the device to be pushed. |
| Speech | OS_186 | No | Because they are not working on actual Speech yet with her. Working on building her mouth muscles. |
| ABA | OS_126 | Yes | It is life changing. With so many therapy hours they can work on communication, negative behaviors, social interaction and also work on any other OT, PT, speech goal. By far the most important and have seen a massive improvement. |
| ABA | OS_143 | Yes | It helped her learn proper behaviors and limit the inappropriate ones. |
| ABA | OS_112 | Yes | The repetitive nature of the therapy helps with building routines which seems to be an effective way to learn skills when there are multi-sensory disorders. |
| ABA | OS_156 | Yes | In my opinion, ABA is most helpful if not necessary before the age of 7. For complex kids like many of ours, it absolutely requires an experienced or gifted/interested practitioner. ABA breaks everything down to small steps and gave my daughter a ton of skills she would not have otherwise. My issue with it is that it works through a transactional relationship and for my daughter's personality, this became and is a problem in her life/our lives. I wish we had started younger and stopped sooner. |
| ABA | OS_152 | Yes | It's a slow progress but she plays more appropriately with toys, follows simple commands, throws her toys less and make her own choices. |
| Occupational | OS_186 | Yes | She has become more outgoing. Making more eye contact. It’s getting easier for us to know what she wants or doesn’t want. She engages with people more than she used to. |
| Occupational | OS_115 | Yes | It has developed her poor fine motor skills by working on finger strength development, isolation, grip, wrist rotation, via structured play tasks and sensory input to arms, wrists and hands. This therapy also works on her self-help and independence skills, for example, moving her hands and her wrists for washing her face and learning to dress/undress, we use the ""backward chaining"" approach for these skills. |
| Occupational | OS_172 | Yes | Has helped with fine motor skills, toileting, dressing, hair brushing, feeding etc. |
| Occupational | OS_177 | No | There haven't been any changes since the beginning of the intervention. |
| Occupational | OS_106 | No | He wasn’t able to understand. |
| Physical | OS_126 | Yes | PT is the only reason why she is walking. Every major milestone was a struggle. [For example] sitting up, standing, etc. Early on her PT pushed her and they were hard therapies to watch because she was crying through each session but it made the world of a difference as she gained strength each session. Aside from the actual sessions, it was very helpful to have their guidance on home support and activities we can do to get her stronger. Also, they suggested devices that will help milestones and get her body ready for walking such as a stander, gait trainer, and AFO's for standing and walking support. |
| Physical | OS_153 | Yes | It has been helpful for choosing and using equipment, particularly for weight bearing (she is non-ambulatory); selecting strollers/wheelchairs; increasing physical activity; getting recommendations for positioning; facilitating fitting of DAFOs (leg braces). |
| Physical | OS_182 | Yes | Before therapy [she] was not able to do anything. Now, she can bring her hands to her mouth, she can turn around (from belly to back without help; from to belly with help), grab toys and play with it! |
| Physical | OS_115 | Yes | I was told [she] would never walk but she did finally take her first steps a few months before her 5th birthday. She required use of a standing frame with leg gaitors for 6o mins almost everyday, use of parallel bars, gait trainers, posture k walkers and AFOS and DAFOS (splints for both feet). She starts to loose tone and some skills if practice isn't kept up (for example she got stiff in certain muscles during Covid time when the input stopped for few months). |
| Physical | OS_122 | Yes | Improved strength, learning to walk, improved core strength, learning to safely move around the house. |
| Physical | OS_148 | No | Sessions were very basic and we found she could complete the skills asked. There was nothing being shown that we weren't already focusing on at home. |

| | | | |
|---|---|---|---|
| Equine | OS_143 | Yes | It helped her ability to focus and overcome some fears; it also helped her core strength and balance. |
| Equine | OS_118 | Yes | [Name] immediately hugs the horse without prompting. She sits perfectly upright and moves accordingly with the horses movement and the increasing speed (not fast but steady increase). She is at her happiest. |
| Equine | OS_119 | Yes | Its calm, controlled and very much about routines. It helps that all the people who attend the stables are quiet but confident too. |
| Equine | OS_112 | No | She did not like being away from the ground and she began having seizures during this time. I might be willing to try it again, but it's more recreational than therapeutic in my opinion unless done in conjunction with an occupational therapist which is normally an extra cost. The traditional horse therapy is normally done by volunteers. |
| Equine | OS_163 | No | We didn't see any difference. It seemed like she liked, or at least tolerated, until we saw that she would cry inconsolably whenever she would watch videos of her session that we took. We stopped the sessions after that. |
| Water | OS_118 | Yes | She is in her element in water and if music playing, she is in heaven. She can stay afloat and can move around happily. |
| Water | OS_152 | Yes | She loves water and enjoys her swim class. She likes to jump in water and go under water. She does not swim independently. It helps with her body awareness, calmness. |
| Water | OS_150 | Yes | She loved it and it makes her happy. |
| Group | OS_137 | Yes | She likes partners, and likes communicating with children. |
| Group | OS_171 | Yes | Helps with interpersonal relationships. |
| Group | OS_177 | No | No changes. |

The OS identification numbers, with the key to identify particular research participants, are only known to the study investigators

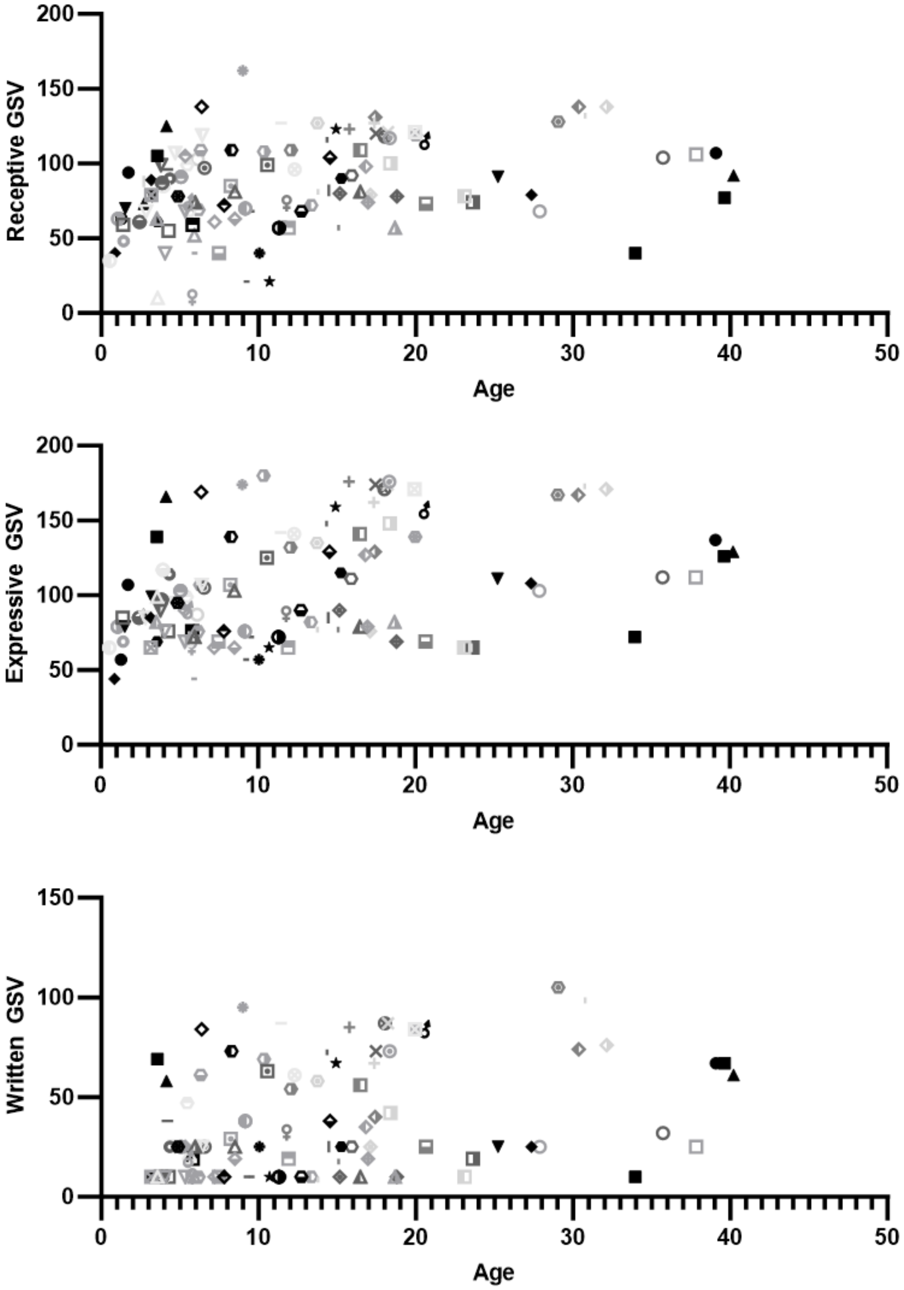


**Supplementary Figure 1a. Communication Subdomain GSV Scores vs Age**

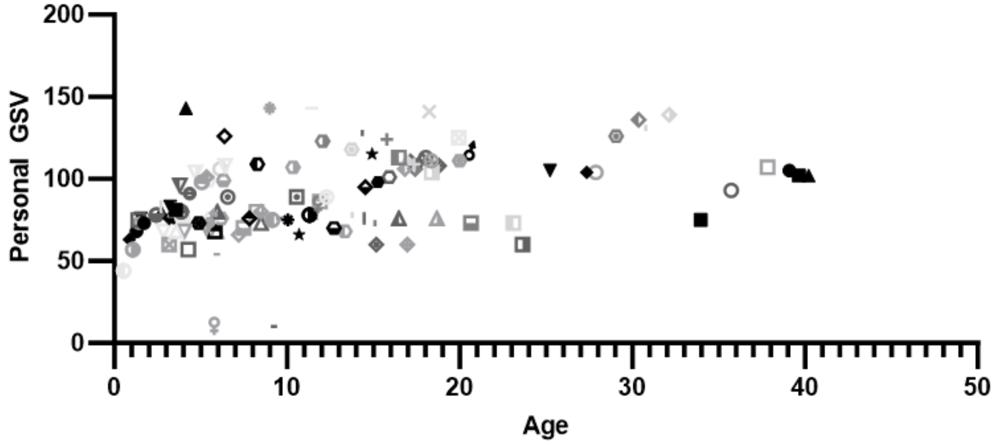


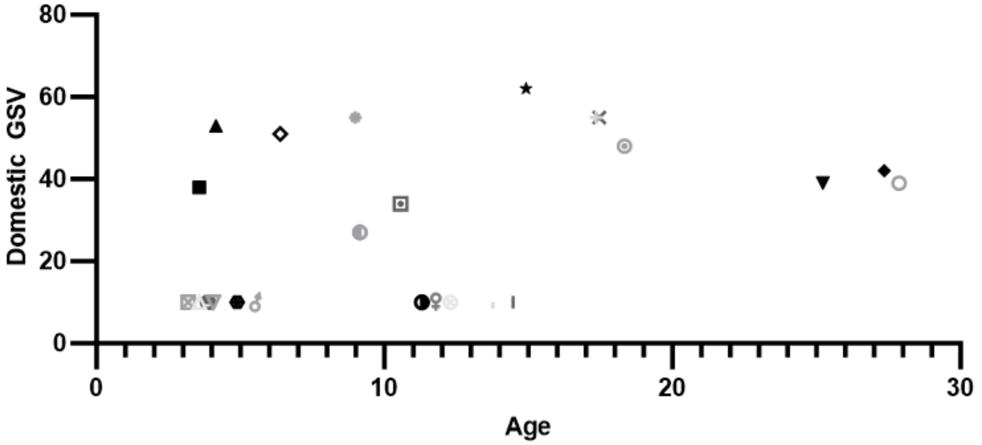


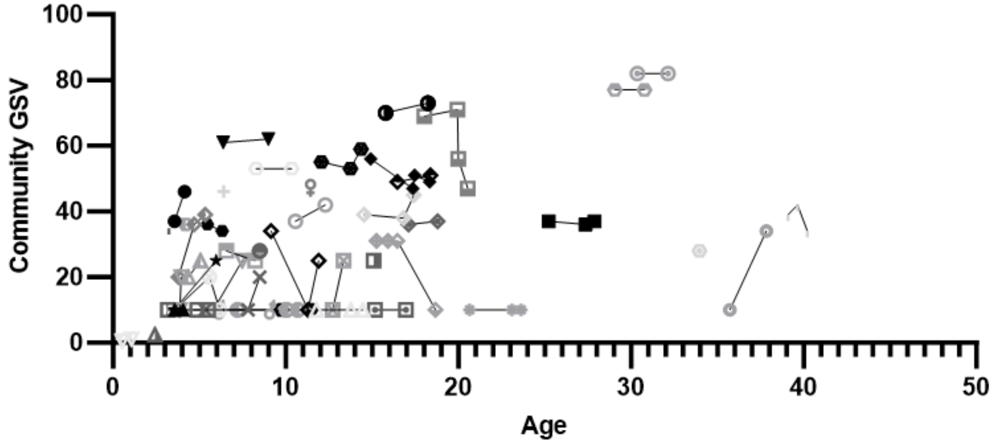


**Supplementary Figure 1b. Daily Living Skills Subdomain GSV Scores vs Age**

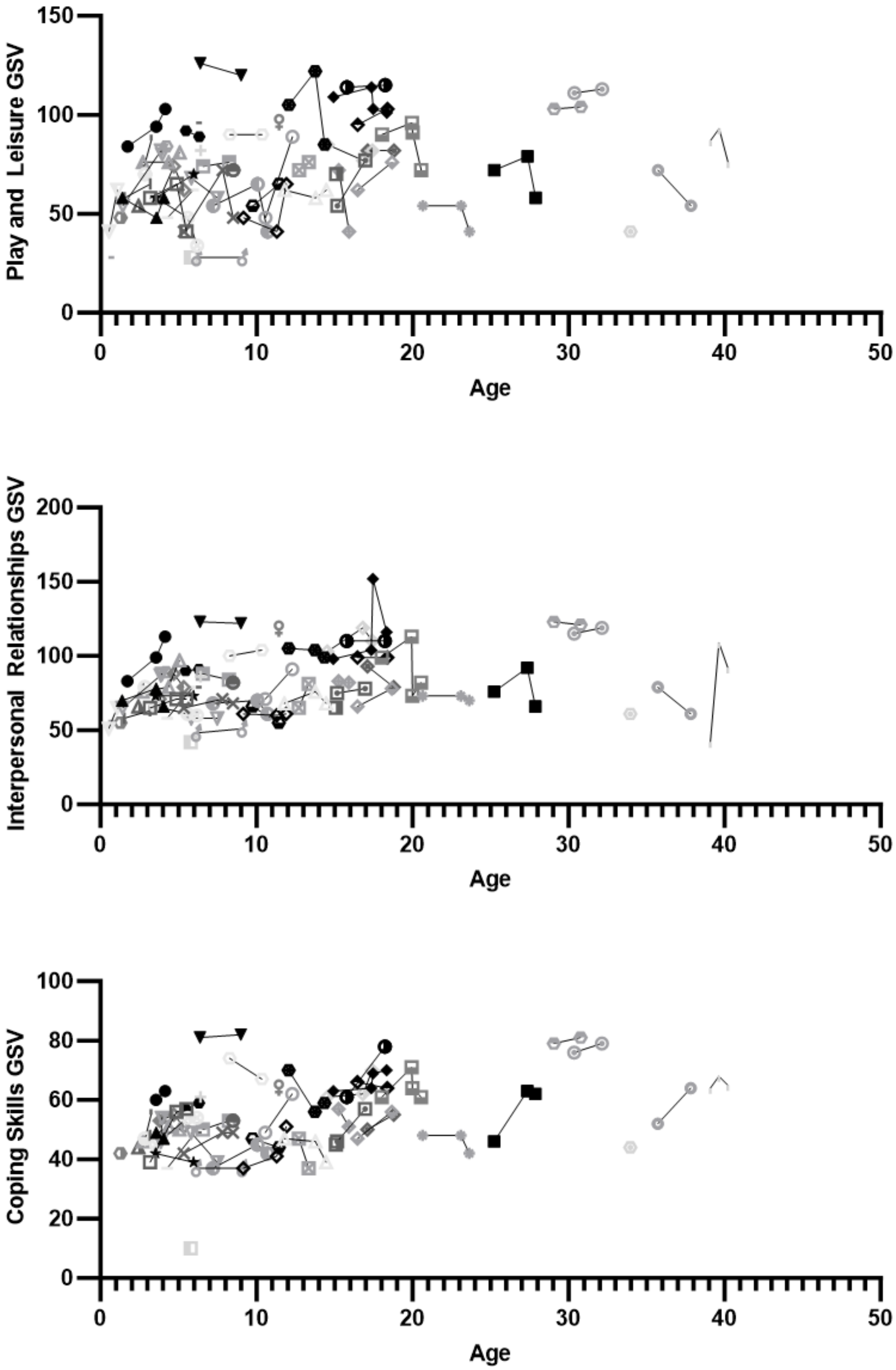


**Supplementary Figure 1c. Socialization Subdomain GSV Scores vs Age**

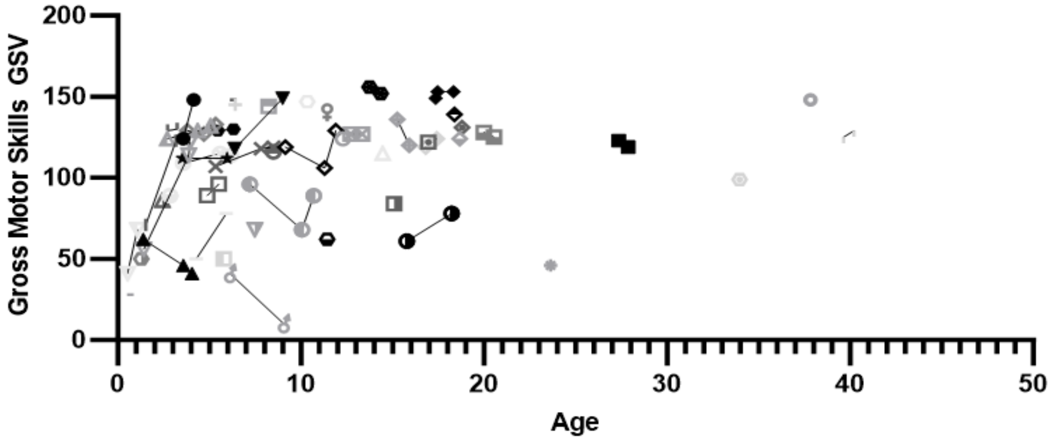


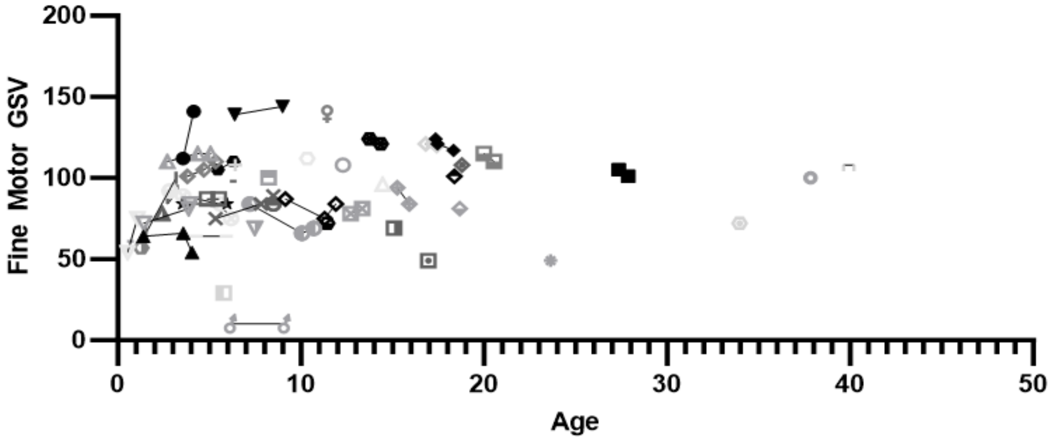


**Supplementary Figure 1d. Motor Subdomain GSV Scores vs Age**

**Supplementary Figure 2. Mutation Counts and Seizure status**

**Supplementary Figure 3. Sub-score analysis of early intervention.** The figure shows the age in months each participant began receiving the therapy, and the sub-score correspondent to the therapy being received. Correlation analysis for all graphs were non-significant.
